# A tool to assess risk of bias in studies estimating the prevalence of mental health disorders (RoB-PrevMH)

**DOI:** 10.1101/2023.02.01.23285335

**Authors:** Thomy Tonia, Diana Buitrago-Garcia, Natalie Peter, Cristina Mesa-Vieira, Tianjing Li, Toshi A. Furukawa, Andrea Cipriani, Stefan Leucht, Nicola Low, Georgia Salanti

**Author notes:** shared first authors.

## Abstract

**Objective:** Biases affect how certain we are about the available evidence, however no standard tool for assessing the risk of bias (RoB) in prevalence studies exists. For the purposes of a living systematic review on prevalence of mental health disorders during the COVID-19 pandemic, we developed a RoB tool to evaluate prevalence studies in mental health (RoB-PrevMH) and tested interrater reliability.

**Methods:** We reviewed existing RoB tools for prevalence studies until September 2020, to develop a tool for prevalence studies in mental health. We tested the reliability of assessments by different users of RoB-PrevMH in 83 studies stemming from two systematic reviews of prevalence studies in mental health. We assessed the interrater agreement by calculating the proportion of agreement and Kappa statistic for each item.

**Results:** RoB-PrevMH consists of three items that address selection bias and information bias. Introductory and signaling questions guide the application of the tool to the review question. The interrater agreement for the three items was 83%, 90% and 93%. The weighted kappa was 0.63 (95% CI 0.54 to 0.73), 0.71 (95% CI 0.67 to 0.85) and 0.32 (95% CI –0.04 to –0.63), respectively.

**Conclusions:** We developed a brief, user friendly, and adaptable tool for assessing RoB in studies on prevalence of mental health disorders. Initial results for interrater agreement were fair to substantial. The tool’s validity, reliability, and applicability should be assessed in future projects.

## 1. Background

Studies of prevalence provide essential information for estimating the burden (1) of mental health conditions, which can inform research and policymaking (2). The pandemic of Coronavirus Disease 2019 (COVID-19), a disease first described in 2020 (3), has generated a large volume of literature rapidly (4), including those on the prevalence of a wide range of conditions, among which those related to mental health.

Increased levels of anxiety, depression, psychological distress, as well as an increase in violent behaviour, alcohol and substance use, among others have been described in association with fear of infection and the effects of contamination measures (1,5). Temporary relief from obligations at school or work, or the need to commute, on the other hand, might alleviate stress for some populations(1).

A systematic review provides a structured way to gather, assess and synthesize evidence from prevalence studies. One essential step in performing a systematic review is the assessment of the risk of bias (RoB) of the included studies (6,7) because the potential biases affect how certain we are about the included evidence and its interpretation (8,9).

There is no agreement on how to assess RoB in prevalence studies (10), despite a ten-fold increase in systematic reviews of prevalence studies in the last decade (11,12). Substantial variability exists in how RoB in prevalence studies have been assessed with more than 30 tools identified and several judged to be inappropriate (13). Notably, some questions/items in existing tools focus on the quality of reporting rather than the RoB.

The purpose of this paper is to present a RoB tool developed to evaluate the risk of bias in studies measuring the prevalence of mental health disorders (RoB-PrevMH). We describe the steps for developing this tool, its items, and the results of interrater agreement obtained by applying the tool to two sets of prevalence studies on mental health disorders.

## 2. Methods

The RoB-PrevMH tool was developed within the MHCOVID project (https://mhcovid.ispm.unibe.ch/), a living systematic review assessing the effect of the COVID-19 pandemic and the containment measures on mental health of the population (1,5,14). MHCOVID involves many volunteers recruited through crowdsourcing to help with data extraction and RoB assessment of a large volume of literature (referred to as the MHCOVID Crowd). We prioritized brevity and ease of application in developing due to the different backgrounds and levels of experience and expertise in the assessment of RoB.

### 2.1 Development of the tool

We searched Medline and Embase (Ovid) from inception to September 2020 to identify published tools or checklists designed to assess the quality, risk of bias, quality of reporting in prevalence studies (Appendix 1). In addition, we searched the Equator network website (https://www.equator-network.org/) and a database of systematic reviews of prevalence studies (15). One reviewer (DBG) screened the search results to identify relevant tools.

We extracted the items from each tool selected for inclusion and grouped them under the domains of selection bias and information bias. For selection bias, items from the existing tools were separated into those referring to population representativeness or to “the proportion of respondents”. For information bias, items from the existing tools were separated into those referring to observer bias, recall bias, or misclassification bias. Items not related to the named biases, were tagged as “other bias” or “reporting”.

Five researchers (DBG, NL, NP, GS, TT) individually went through the list of questions in each included tool, excluded duplicated questions, and marked those that were most relevant for prevalence studies for mental health disorders. They then discussed their assessments and reached consensus prior to drafting the first version of the tool and the signaling question. Figure 1 illustrates the process of developing RoB-PrevMH.

**Figure 1:**
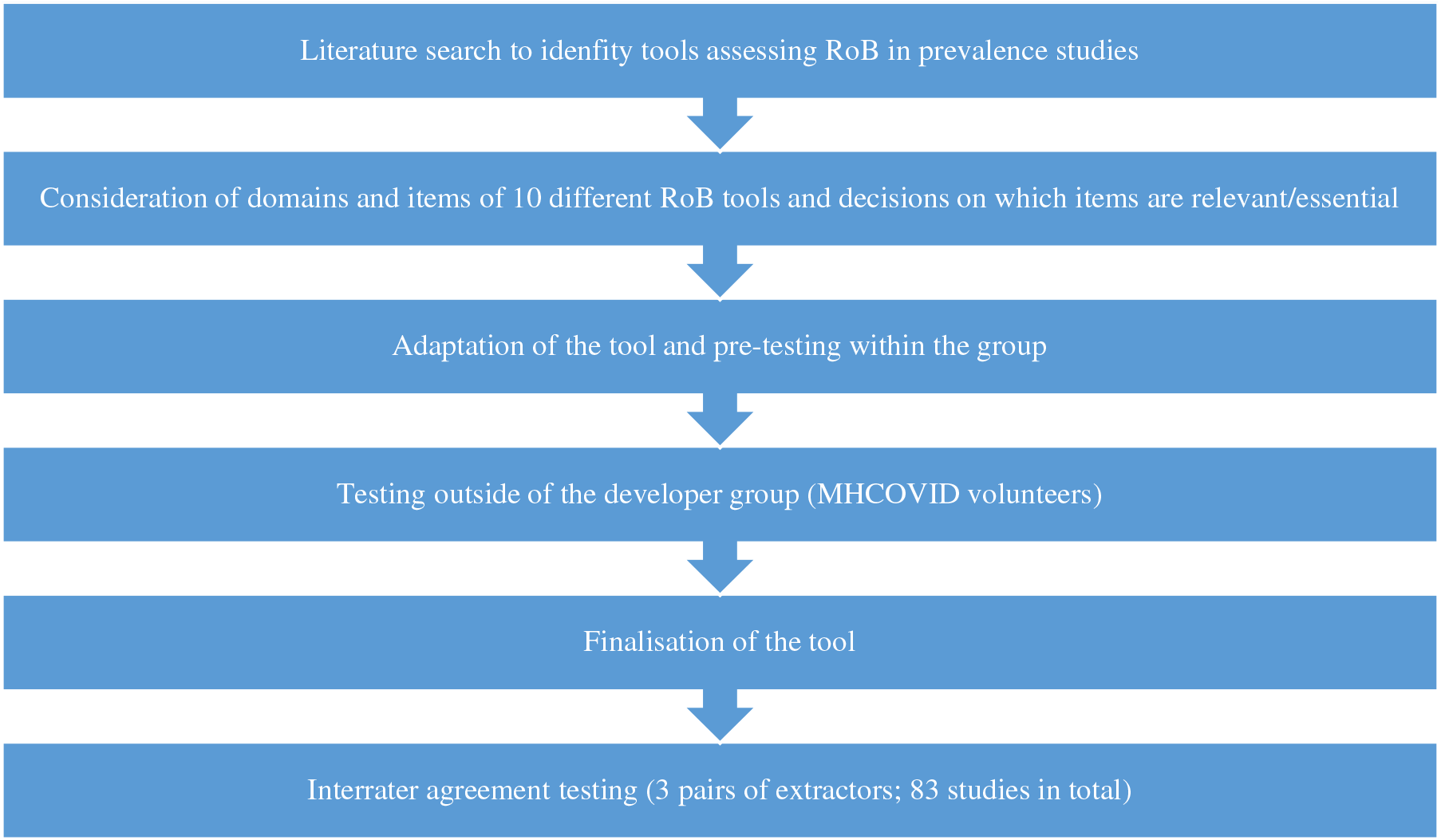
Process of developing and testing the RoB-PrevMH.

### 2.2 Testing and finalization of the tool

Four members of the team (SL, NP, GS, TT) pilot tested the first version of the tool and drafted a guidance document. Subsequently, these four researchers and four volunteers from the MHCOVID Crowd (who were not involved in the development of the tool) further tested the tool in a total of 8 studies. Based on feedback from this exercise the guidance document was updated accordingly, including examples and practical advice.

### 2.3 Interrater reliability

We tested the reliability of assessments by different users of RoB-PrevMH with two sets of prevalence studies.

The first set included 50 prevalence studies (two sets of 25) randomly selected from those identified as potentially relevant for the MHCOVID project during the abstract screening stage. Two pairs of researchers independently applied RoB-PrevMH (team A, 25 studies: CMV and TT; team B 25 studies: DBG and NP). The second set included 33 studies from a systematic review of the prevalence of post-traumatic stress disorders, major depressive disorder, and generalized anxiety disorder in migrants with pre-migration exposure to armed conflict (16). By using this second set of studies, we examined how RoB-PrevMH performed in a research question that was different from the one it was originally developed for. Two researchers (team C: DBG and CMV) independently applied RoB-PrevMH in this set of studies.

To assess the extent of reproducibility we calculated the unweighted and weighted kappa statistic (with 95% confidence intervals [CI]). To take into consideration partial agreement in each item of the tool, a weight of 1 indicated perfect agreement, 0.5 for partial agreement and 0 for complete disagreement (17,18). The analysis was conducted in STATA 15.1 (19), as well as the percentage of agreement between raters (number of agreements/number of assessments × 100). We followed the interpretation of the Kappa statistic proposed by Landis and Koch (1977) and described in the STATA manual (18) where the values below the cut points 0.00, 0.20, 0.40, 0.60, 0.80 and 1.00 approximately define poor, slight, fair, moderate substantial and almost perfect agreement (20).

## 3. Results

### 3.1 Description of RoB-PrevMH tool

We identified ten tools that assess RoB in prevalence studies, summarized in Table 1 (21–30). Following the process mentioned above, we developed the RoB-PrevMH which consists of one introductory question and three items (Table 2a). It also includes signaling questions aimed to help the user reach a judgement (31). The elaboration and guidance document are presented in Appendix 2. The risk of bias for each item can be judged as “high”, “low”, or “unclear”. When possible, we instructed users to avoid judging any of the questions as unclear, unless necessary. The tool does not allow a summary RoB assessment because it has been suggested that some aspects of study quality might be more important than aggregated scores and that such scores are problematic. (32,33)

**Table1:** RoB tools considered for developing RoB-PrevMH.

| ID | Tool | Description | Number of items/questions | Validation process |
| --- | --- | --- | --- | --- |
| 1 | Leboeuf-Yde, et al 1995 | A tool designed to assess the quality of prevalence studies on low back pain. | Eleven methodologic criteria | Not reported |
| 2 | Loney P, et al 1998 | A critical appraisal tool designed to assess the methodological strengths, results and relevance of articles on prevalence of a health problem. | Eight items with a scoring system | Consensus between two assessors |
| 3 | Boyle M, et al 1998 | A guideline to critically appraise prevalence studies on psychiatric disorders, both in the general population and in specific clinical settings | Evaluates three main items divided in 11 questions | Not reported |
| 4 | Silva L, et al 2001 | A tool to assess the usefulness of prevalence studies in the context of surveillance activities | Covers six technical aspects divided in 19 questions with a scoring system | Consensus for the scoring system |
| 5 | Shamliyan T, et al 2010 | A tool for evaluating the quality of studies that examine the prevalence of chronic conditions or risk factors. | Six criteria for external validity and five for internal validity | The tool was tested in four studies of incidence or prevalence. Kappa values showed fair agreement. |
| 6 | Hoy D et al, 2012 | A risk of bias tool for prevalence studies based on Leboeuf-Y de et al 1995 | Ten items plus a summary assessment. | Overall interrater agreement=91%<br>Kappa= 0.82 (95% CI: 0.76, 0.86) |
| 7 | Giannakopoulos D et al. 2012 | An instrument for the qualitative assessment of the methodology of prevalence studies | Ten items with a scoring system | **Pilot phase**<br>Kappa for the quality score= mean 0.62 ±0.15<br>Kappa for individual questions= mean 0.78±0.27<br>**After feedback**<br>Kappa for the quality score= range 0.94-1.00 |
| 8 | Munn Z et al 2014 | A critical appraisal tool for assessing studies included in systematic reviews of prevalence. | Ten questions | 5-point Likert scale (1 very unacceptable, 5 very acceptable)<br>Ease of tool use= mean 3.63 ±0.72<br>Acceptability= mean 4.33 ±0.49<br>Timeliness= mean 3.94 ±0.57 |
| 9 | The Joanna Briggs Institute. 2016 | A tool to assess the methodological quality of a prevalence study and the possibility of bias. | Nine questions with an overall appraisal question. | Not reported |
| 10 | Pega F et al 2019 | A tool for assessing the risk of bias in prevalence studies of exposure to occupational risk factors. | Eight domains | Using a raw measure of agreement, the tool achieved substantial agreement in six domains (conflict of interest, other bias, lack of blinding of study personnel, exposure misclassification, selective reporting of exposures) and poor agreement in two domains (incomplete exposure data, selection of participants into the study). |

**Table 2a:**
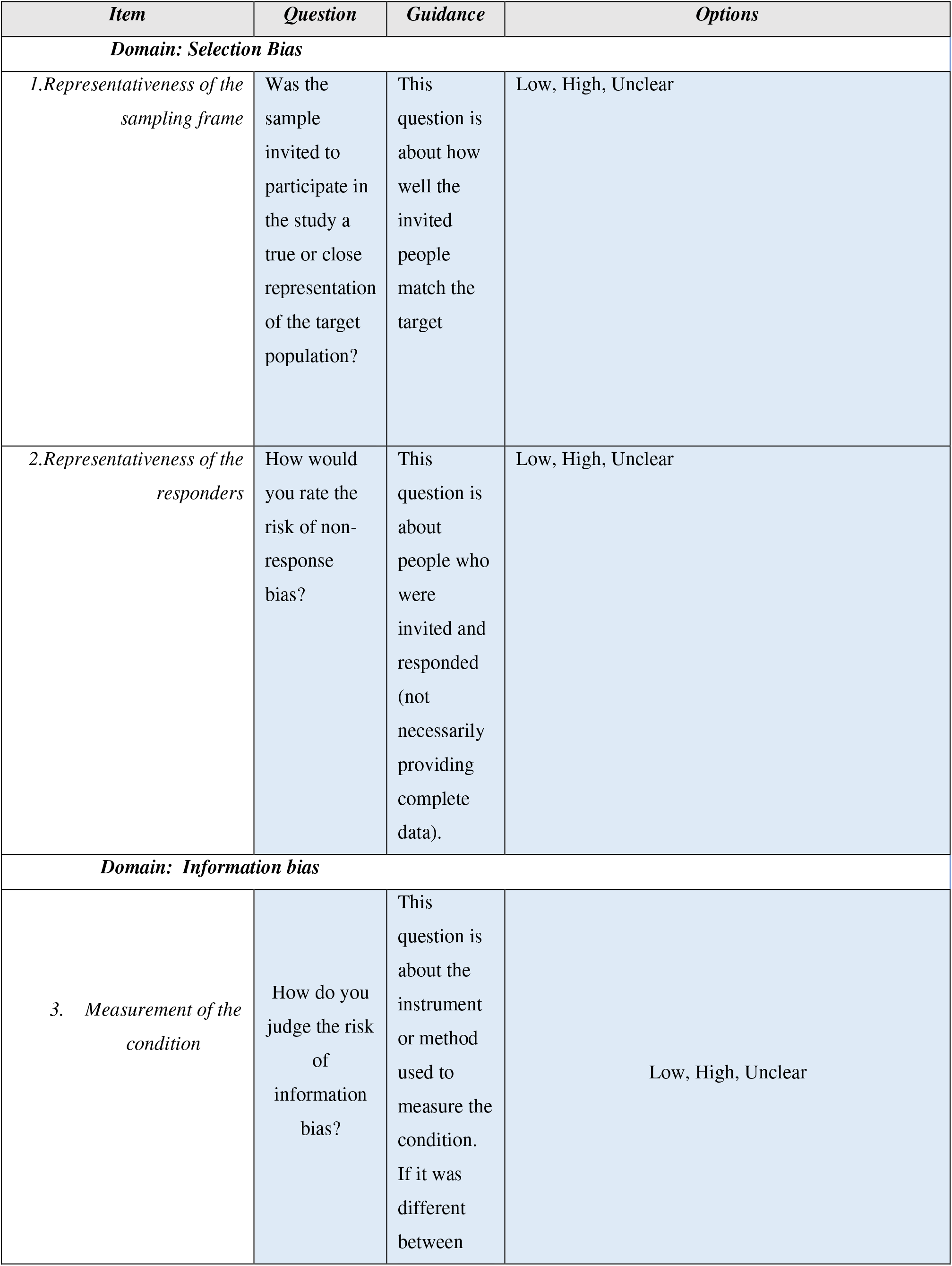
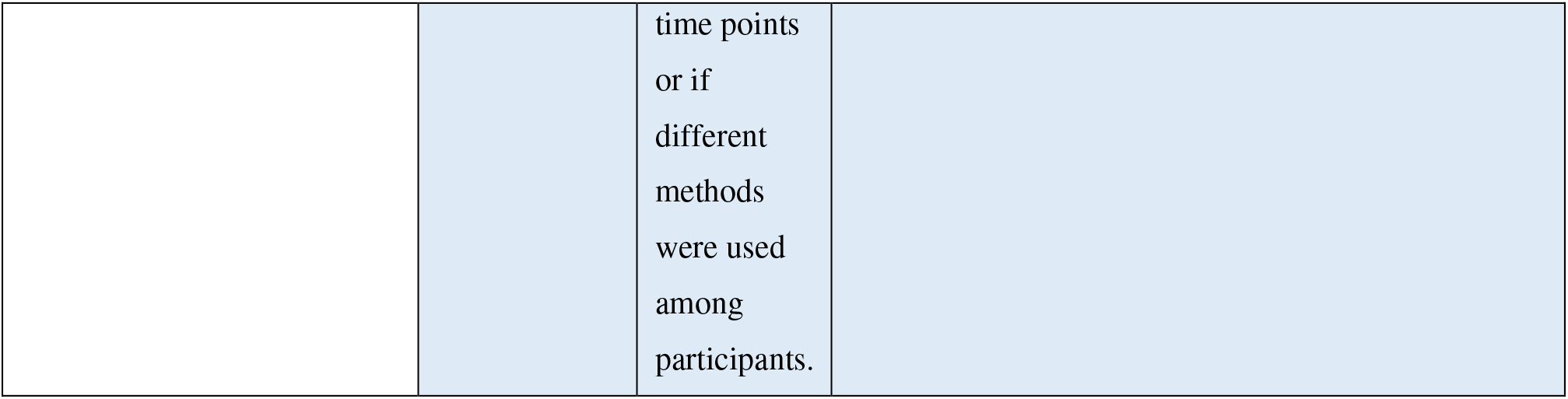
Items included in the RoB-PrevMH.

**Table 2b:** Suggested changes in the phrasing of the RoB-RrevMH items.

| *Item* | *Question* | *Guidance* | *Options* |
| --- | --- | --- | --- |
| **Domain: Selection Bias** |  |  |  |
| *1.Representativeness of the sampling frame* | Was the sample invited to participate in the study a true or close representation of the target population? | This question is about how well the invited people match the target (general) population | Low, High, Unclear |
| *2.Representativeness of the responders* | Was the sample that provided data a true or close representation the sample invited to participate? | This question is about people who were invited and responded (not necessarily providing complete data). | Low, High, Unclear |
| **Domain: Information bias** |  |  |  |
| *3. Measurement of the condition* | Was the condition measured/detected in an unbiased and reproducible way? | This question is about the instrument or method used to measure the condition. If it was different between time points or if different methods were used among participants. | Low, High, Unclear |
|  | - Was the tool used to measure the condition validated? *<br>Were the methods for measuring the condition | Observer bias might occur when the training of observers was not done or the procedures to collect | Low, High, Unclear |
|  | standardised?<br>- Does the measurement of the condition depend on the memory of the participants? | the data are not the same for every participant included in the study.<br>Recall bias might occur when questions refer to the past and their answering depends on the condition |  |

\* this was a requirement for inclusion in the MHCovid study and hence not included in the current version of the tool

The introductory question is “*Was the target population clearly defined?*”. By “*target population*” we refer to the entire population for which we are interested to draw inference. In the first set of studies from the MHCOVID project, the target population of the systematic review was defined as “the general population” or any age or gender-based subgroups of the general population (e.g., children only, or men only, or elderly, see Appendix). In the second set of studies, the target population of the systematic review was migrants exposed to armed conflict (16).

This introductory question had two response options; “yes” or “no” and has implications for the evaluation of the first RoB item: if the answer is “no”, the first item of the tool is automatically assigned an “unclear” risk.

Item 1 Selection bias: Representativeness of the sampling frame

The first RoB item is related to the representativeness of the sample invited with respect to the target population by asking whether “Was *the sample invited to participate in the study a true or close representation of the target population?*” The signaling question for this item asked about the method for recruitment of participants and based on the response, the instructions guided the user to reach the corresponding RoB judgement (for example low risk when the total or a randomly selected sample of the target population was invited; high risk for open calls for participation online, or quota sampling; and unclear risk when the method to invite participants and the specific context of the sampling was not specified or when the target population was not defined; for more details see the instructions in the Supplement).

Item 2 Selection bias: Representativeness of the responders

The second item requires a judgement as to whether those who declined the invitation, in relation to those who participated in the study, would introduce bias in the prevalence estimate, “*How would you rate the risk of non-response bias*?” The reasons of non-participation are instrumental in forming a judgement about the risk of bias. However, these are rarely reported, if ever. We assumed that in our context the decision not to participate is associated, directly or indirectly with the mental health of the persons invited to the study. The signaling question for this item inquiries therefore only about the proportion of participants providing data out of the number of people invited to participate and based on the response, the RoB judgement is made.

Item 3 Information bias: Measurement of the condition

The last item assesses the likelihood of misclassification of the methods used to measure the target condition, *“How do you judge the risk of information bias*?”. We provide guidance for judging this question for the MHCOVID project (Appendix 2); for instance, if the tool/method used to measure the condition was not applied properly across time points or across groups of participants.

### 3.2 Interrater agreement

Table 3 shows the results of the interrater agreement for each item of the RoB-PrevMH, including both weighted and unweighted Kappa for the 83 included studies. For item 1, the interrater agreement was substantial with weighted Kappa=0.63 (95% CI 0.54 to 0.73) and overall agreement 75%. For item 2, the agreement was substantial, with weighted Kappa statistic equal to 0.71 (95% CI 0.67 to 0.85) and overall agreement 82%. For item 3, the kappa was 0.32 (95% CI -0.04 to 0.63; overall agreement 89%), classifying interrater agreement as fair.

**Table 3:** Results of interrater agreement testing.

| Item | Unweighted Kappa (95% CI) | % Agreement | Weighted Kappa (95% CI) | % Agreement |
| --- | --- | --- | --- | --- |
| 1.Representativeness of the sampling frame | 0.60 (0.48-0.68) | 74.7 | 0.63 (0.54 - 0.73) | 83.1 |
| 2.Representativeness of the responders | 0.69 (0.59-0.70) | 81.9 | 0.71 (0.67 - 0.85) | 90.3 |
| 3. Measurement of the condition | 0.28 (0.10-0.73) | 89.2 | 0.32 (-0.04 -0.63) | 93.4 |

There was a total of 45 disagreements out of 249 paired assessments among 83 studies. Most of the disagreements (n=35) were between “unclear” and either “high” or “low”. Ten disagreements were between “high” versus “low” assessments.

## 4. Discussion

### 4.1 Summary of findings

We developed the RoB-PrevMH, a concise risk of bias tool for prevalence studies in mental health that was designed with the intention to be adaptable to different systematic reviews and consisting of three items: representativeness of the sample, non-response bias, and information bias. Our tool showed fair to substantial interrater reliability when applied to studies included in two systematic reviews of prevalence studies. All three items from RoB-PrevMH have been considered or included in existing tools (22,26,29). RoB-PrevMH does not contain any item on reporting and does not require an assessment of the overall RoB in a study. For each item, three assessments of risk of bias are possible (high, unclear and low)

### 4.2 Strengths and limitations

The strengths of RoB-PrevMH include the fact that it was created after a comprehensive review of items identified in previous tools as well as a consensus between researchers. Second, the feedback we received from the MH-COVID Crowd who used the tool suggests that the tool is concise and easy to use. In addition, it focuses on RoB only and avoids questions that assess reporting. Given the small number of items, RoB-PrevMH probably requires less time than other published tools, which is important when evidence is accumulating rapidly and needs to be assessed and incorporated into systematic reviews as soon as possible. Furthermore, the tool was tested by three pairs of extractors in two sets of studies with different aims. The interrater reliability was rated from fair to substantial. Finally, the tool has the potential to be tailored to other research questions.

Our tool also has limitations. We developed the tool after discussion and testing among rather small teams of methodologists and investigators. A wider consultation strategy that involved public mental health experts, investigators that have designed and undertaken many prevalence studies and more methodologists could have delivered a more refined and more acceptable tool. Then, the brevity of the tool could be considered a potential limitation. For example, the MHCOVID project only includes studies that used validated tools for measuring mental health outcomes, so we did not include specific items for recall bias and observer bias, which might be important for other questions. Even if we assume that it would be quicker to complete than other tools due to its brevity, we did not formally assess the time required for completion in comparison to other tools. Additionally, the need to tailor the tool for each project and create training material for the people who will apply it accordingly, might require more time upon initiation of a project compared to other tools. Moreover, the interrater reliability varied between the three items, with kappa values ranging from 0.32 to 0.71.

Even though representativeness might be difficult to judge objectively, the interrater agreement for this item was substantial and is higher than the tool by Hoy (26), that assessed the interrater agreement in 54 studies. For the second item on non-response, interrater agreement was substantial, but lower than similar items in the Hoy tool (26). The third item on misclassification was found to have the lowest kappa but the highest agreement between raters. In classification tables with great imbalance in the marginal probabilities and a high underlying correct classification rate kappa can be paradoxically low, as it is the case of kappa for information bias (34,35). We do not make an overall RoB assessment for each study like the Hoy tool does (26), since it has been shows that this is problematic(32).

### 4.3 Application of RoB-PrevMH in future projects

The design of prevalence studies differs substantially depending on the question they intend to answer; as a result, having a universal tool for all types of prevalence studies, like we have for RCTs and some observational studies, (36–38) might not be realistic; instead, we need tools that can be tailored to specific research questions (39).

Future projects applying RoB-PrevMH might need to modify the questions (Table 2b provides suggestions to improve the phrasing of the questions). The questions can also be tailored to specific review questions of future projects. RoB-PrevMH was conceptualized and developed for the MHCOVID project (5). An inclusion criterion required the use of a validated assessment tool. For this reason, RoB-PrevMH does not consider the use of validated tools in the information bias domain. This may not be the case for a future project in which validated diagnostic tools do not exist for a condition (for instance cognitive deficits in post-COVID-19) or the project does not impose inclusion criterion such as ours. Another example comes from the MHCOVID project itself. In this project we decided to rate the risk of bias for the second and third item at every follow up time point instead of following the original instructions to give one global rating for each study. Other projects might consider the idea of not having an arbitrary threshold for the proportion of respondents and instead extract the reported proportion and analyze the data by conducting pre-specified subgroup analyses, based on this continuum of response rate with meta-regression. Moreover, our chosen arbitrary threshold for response rate might be inappropriate for other studies, as we included studies on the general population, during a pandemic and mostly done online; in other settings a “good” response rate might be higher than 70%. Training for the tool should be tailored to a specific project and include relevant examples. For instance, for the MHCOVID project, we developed an educational video and provided online training for the volunteers of the project who extracted data from included studies and conducted the RoB assessment (https://mhcovid.ispm.unibe.ch/crowd.html).

Assessment of the risk of bias in prevalence studies applies to any condition. The tools that have been published were mostly developed for specific situations, ranging from low back pain to exposure to occupational risk factors. The methods that we used to develop RoB-PrevMH follow recommended methods for the development of guidelines (40)and should be used to further develop a risk of bias tool that can be applied to any systematic review question that aims to summarise the prevalence of a condition or risk factor. The MHCOVID project has provided the basis for building a network or experts with experience of RoB assessment (36,38) and critical appraisal of prevalence studies (13,28) to develop a generic framework for tools to assess the RoB in prevalence studies (41).

## Conclusion

RoB-PrevMH is a brief and adaptable tool for assessing RoB in studies on prevalence of mental health disorders. Initial results for interrater agreement were fair to substantial. The validity, reliability and applicability of RoB-PrevMH should be further assessed in future projects.

## Data Availability

All data produced in the present study are available upon reasonable request to the authors

## Acknowledgements

The authors acknowledge the contribution of Anna Ceraso, Aoife O’ Mahony, Trevor Thompson and Marialena Trivella who tested and gave feedback on the tool; the contribution of Alexander Holloway for technical support and the contribution of Leila Darwish for her support in the MHCOVID project.

## Contributors

GS, NL, TL, TT, NP, DBG, CMV, TWF, AC, SL designed the study; TT, DBG, NP, CVM collected data; TF, GS, TT performed the statistical analysis; first draft was prepared by TT and DBG; revised and approved by all

## References

1. Leucht S, Cipriani A, Furukawa TA, Peter N, Tonia T, Papakonstantinou T, et al. A living meta-ecological study of the consequences of the COVID-19 pandemic on mental health. Eur Arch Psychiatry Clin Neurosci. 2021 Mar;271(2):219–21.

2. Harder T. Some notes on critical appraisal of prevalence studies: Comment on: ‘The development of a critical appraisal tool for use in systematic reviews addressing questions of prevalence’. Int J Health Policy Manag. 2014 Oct;3(5):289–90.

3. Liu YC, Kuo RL, Shih SR. COVID-19: The first documented coronavirus pandemic in history. Biomed J. 2020 Aug;43(4):328–33.

4. Ipekci AM, Buitrago-Garcia D, Meili KW, Krauer F, Prajapati N, Thapa S, et al. Outbreaks of publications about emerging infectious diseases: the case of SARS-CoV-2 and Zika virus. BMC Med Res Methodol. 2021 Dec;21(1):50.

5. Salanti G, Peter N, Tonia T, Holloway A, White IR, Darwish L, et al. The Impact of the COVID-19 Pandemic and Associated Control Measures on the Mental Health of the General Population: A Systematic Review and Dose–Response Meta-analysis. Ann Intern Med. 2022 Nov;175(11):1560–71.

6. Katikireddi SV, Egan M, Petticrew M. How do systematic reviews incorporate risk of bias assessments into the synthesis of evidence? A methodological study. J Epidemiol Community Health. 2015 Feb;69(2):189–95.

7. Systematic Reviews in Health Research: Meta-Analysis in Context, 3rd Edition | Wiley [Internet]. Wiley.com. 2022 [cited 2022 Jun 29]. Available from: https://www.wiley.com/en-us/Systematic+Reviews+in+Health+Research%3A+Meta+Analysis+in+Context%2C+3rd+Edition-p-9781405160506

8. Guyatt GH, Oxman AD, Vist G, Kunz R, Brozek J, Alonso-Coello P, et al. GRADE guidelines: 4. Rating the quality of evidence--study limitations (risk of bias). J Clin Epidemiol. 2011 Apr;64(4):407–15.

9. Viswanathan M, Berkman ND, Dryden DM, Hartling L. Assessing Risk of Bias and Confounding in Observational Studies of Interventions or Exposures: Further Development of the RTI Item Bank [Internet]. Rockville (MD): Agency for Healthcare Research and Quality (US); 2013 [cited 2022 Jun 29]. (AHRQ Methods for Effective Health Care). Available from: http://www.ncbi.nlm.nih.gov/books/NBK154461/

10. Borges Migliavaca C, Stein C, Colpani V, Barker TH, Munn Z, Falavigna M, et al. How are systematic reviews of prevalence conducted? A methodological study. BMC Med Res Methodol. 2020 Apr 26;20(1):96.

11. Migliavaca CB, Stein C, Colpani V, Barker TH, Ziegelmann PK, Munn Z, et al. Meta-analysis of prevalence: I 2 statistic and how to deal with heterogeneity. Res Synth Methods. 2022 May;13(3):363–7.

12. Hoffmann F, Eggers D, Pieper D, Zeeb H, Allers K. An observational study found large methodological heterogeneity in systematic reviews addressing prevalence and cumulative incidence. J Clin Epidemiol. 2020 Mar;119:92–9.

13. Migliavaca CB, Stein C, Colpani V, Munn Z, Falavigna M, Prevalence Estimates Reviews – Systematic Review Methodology Group (PERSyst). Quality assessment of prevalence studies: a systematic review. J Clin Epidemiol. 2020 Nov;127:59–68.

14. Salanti G, Cipriani A, Furukawa TA, Peter N, Tonia T, Papakonstantinou T, et al. An efficient way to assess the effect of COVID-19 on mental health in the general population. Lancet Psychiatry. 2021 May;8(5):e14–5.

15. Buitrago-Garcia D. Meta-análisis de prevalencia: Revisión sistemática de los métodos utilizados, propuesta de una herramienta para evaluar la calidad y evaluación de los diferentes métodos estadísticos utilizados para meta analizar prevalencias. Vol. Magister en Epidemiología Clínica, Facultad de Medicina. [Bogotá]: Universidad Nacional de Colombia; 2018.

16. Mesa-Vieira C, Haas AD, Buitrago-Garcia D, Roa-Diaz ZM, Minder B, Gamba M, et al. Mental health of migrants with pre-migration exposure to armed conflict: a systematic review and meta-analysis. Lancet Public Health. 2022 May;7(5):e469–81.

17. Kirkwood BR, Sterne JAC, Kirkwood BR. Essential medical statistics. 2nd ed. Malden, Mass: Blackwell Science; 2003. 501 p.

18. StataCorp. Stata 17 Base Reference Manual. College Station, TX: Stata Press; 2017.

19. McHugh ML. Interrater reliability: the kappa statistic. Biochem Medica. 2012;22(3):276–82.

20. Landis JR, Koch GG. The Measurement of Observer Agreement for Categorical Data. Biometrics. 1977 Mar;33(1):159.

21. Leboeuf-Yde C, Lauritsen JM. The prevalence of low back pain in the literature. A structured review of 26 Nordic studies from 1954 to 1993. Spine. 1995 Oct 1;20(19):2112–8.

22. Loney PL, Chambers LW, Bennett KJ, Roberts JG, Stratford PW. Critical appraisal of the health research literature: prevalence or incidence of a health problem. Chronic Dis Can. 1998;19(4):170–6.

23. Boyle MH. Guidelines for evaluating prevalence studies. Evid Based Ment Health. 1998 May 1;1(2):37–9.

24. Silva LC, Ordúñez P, Paz Rodríguez M, Robles S. A tool for assessing the usefulness of prevalence studies done for surveillance purposes: the example of hypertension. Rev Panam Salud Publica Pan Am J Public Health. 2001 Sep;10(3):152–60.

25. Shamliyan TA, Kane RL, Ansari MT, Raman G, Berkman ND, Grant M, et al. Development quality criteria to evaluate nontherapeutic studies of incidence, prevalence, or risk factors of chronic diseases: pilot study of new checklists. J Clin Epidemiol. 2011 Jun 1;64(6):637–57.

26. Hoy D, Brooks P, Woolf A, Blyth F, March L, Bain C, et al. Assessing risk of bias in prevalence studies: modification of an existing tool and evidence of interrater agreement. J Clin Epidemiol. 2012 Sep;65(9):934–9.

27. Giannakopoulos NN, Rammelsberg P, Eberhard L, Schmitter M. A new instrument for assessing the quality of studies on prevalence. Clin Oral Investig. 2012 Jun;16(3):781–8.

28. Munn Z, Moola S, Riitano D, Lisy K. The development of a critical appraisal tool for use in systematic reviews addressing questions of prevalence. Int J Health Policy Manag. 2014 Aug 13;3(3):123–8.

29. Joanna Briggs Institute. The Joanna Briggs Institute Critical Appraisal tools for use in JBI Systematic Reviews Checklist for Prevalence Studies [Internet]. Joanna Briggs Institute; 2017. Available from: https://jbi.global/sites/default/files/2019-05/JBI_Critical_Appraisal-Checklist_for_Prevalence_Studies2017_0.pdf

30. Pega F, Norris SL, Backes C, Bero LA, Descatha A, Gagliardi D, et al. RoB-SPEO: A tool for assessing risk of bias in studies estimating the prevalence of exposure to occupational risk factors from the WHO/ILO Joint Estimates of the Work-related Burden of Disease and Injury. Environ Int. 2020 Feb;135:105039.

31. Morgan RL, Thayer KA, Santesso N, Holloway AC, Blain R, Eftim SE, et al. A risk of bias instrument for non-randomized studies of exposures: A users’ guide to its application in the context of GRADE. Environ Int. 2019 Jan;122:168–84.

32. Jüni P. The Hazards of Scoring the Quality of Clinical Trials for Meta-analysis. JAMA. 1999 Sep 15;282(11):1054.

33. Stroup DF, Berlin JA, Morton SC, Olkin I, Williamson GD, Rennie D, et al. Meta-analysis of observational studies in epidemiology: a proposal for reporting. Meta-analysis Of Observational Studies in Epidemiology (MOOSE) group. JAMA. 2000 Apr 19;283(15):2008– 12.

34. Cicchetti DV, Feinstein AR. High agreement but low kappa: II. Resolving the paradoxes. J Clin Epidemiol. 1990 Jan;43(6):551–8.

35. Feinstein AR, Cicchetti DV. High agreement but low Kappa: I. the problems of two paradoxes. J Clin Epidemiol. 1990 Jan;43(6):543–9.

36. Higgins JPT, Altman DG, Gotzsche PC, Juni P, Moher D, Oxman AD, et al. The Cochrane Collaboration’ s tool for assessing risk of bias in randomised trials. BMJ. 2011 Oct 18;343(oct18 2):d5928–d5928.

37. Jørgensen L, Paludan-Müller AS, Laursen DRT, Savović J, Boutron I, Sterne JAC, et al. Evaluation of the Cochrane tool for assessing risk of bias in randomized clinical trials: overview of published comments and analysis of user practice in Cochrane and non-Cochrane reviews. Syst Rev. 2016 May 10;5:80.

38. Sterne JA, Hernán MA, Reeves BC, Savović J, Berkman ND, Viswanathan M, et al. ROBINS-I: a tool for assessing risk of bias in non-randomised studies of interventions. BMJ. 2016 Oct 12;i4919.

39. Buitrago-Garcia D, Salanti G, Low N. Studies of prevalence: how a basic epidemiology concept has gained recognition in the COVID-19 pandemic. BMJ Open. 2022 Oct 1;12(10):e061497.

40. Moher D, Schulz KF, Simera I, Altman DG. Guidance for Developers of Health Research Reporting Guidelines. PLoS Med. 2010 Feb 16;7(2):e1000217.

41. Buitrago-Garcia, D. Development of a Risk of Bias tool for prevalence studies. [Internet]. 2023. Available from: https://osf.io/b4qt9

