## Appendix 1 for "A tool to assess risk of bias in studies estimating the prevalence of mental health disorders (RoB-PrevMH)"

Appendix X. Search strategy for identifying tools to assess the risk of bias in prevalence studies

1 exp Prevalence/
2 prevalence.ti,ab.
3 prevalence stud*.ti,ab.
4 or/1-3
5 risk of bias.ti,ab.
6 exp Bias/
7 bias.ti,ab.
8 methodological quality.ti,ab
9 or/5-8
10 exp Checklist/
11 checklis*.ti,ab.
12 assessment tool.ti,ab.
13 exp Guideline/
14 guidelin*.ti,ab.
15 or/9-13
16 4 and 14 and 15
