## Appendix 2 for "A tool to assess risk of bias in studies estimating the prevalence of mental health disorders (RoB-PrevMH)"

### RISK OF BIAS TOOL AND INSTRUCTIONS

| | |
|---|---|
| **Question:**<br>Was the target population clearly defined? | This question refers to *the population we want to know about (the one that you chose as your population in the tagging section of the data extraction form)*, not the sample that was collected.<br>✓ **Yes,** if<br>• the target population is described as “the general population” or any age or sex based subgroups of the general population (e.g. the target population is children only, or men only, or elderly only)<br>○ we also accept descriptions that are indicative of the general population (but don’t mention the exact term) like “people in India”/”adults in Greece”/”eligible were all people residing in the US”<br>• the target population is a specific population sufficiently defined in terms of a) geographical location b) important characteristics such as comorbidities, occupation, living situation etc.<br>**Example**: “... pregnant women in Italy”, “... college students”).<br>✗ **No,** if important characteristics are missing in the description of the target population or no information is given<br><br>If the question is answered “yes”, type (or copy-paste) the definition of the target population in the field and move to the next RoB question. If the specific geographical region is not stated (but it is a general population and or a described specific population), add in the description the country where the study has taken place.<br>If this question is answered “no”, the next question does not appear, but is answered with “unclear risk” automatically |
| **RoB Question 1:**<br>Was the sample invited to participate in the study a true or close representation of the target population? | This question is about the *people who were invited* and how well this matches the target population.<br><br>✗ **High risk** if the methods used to identify the participants are likely to produce a sample that is different to the target population in important characteristics (such as sex, age, comorbidities, socioeconomical status etc).<br>Some of the common methods that will produce a non-representative sample are<br>• open call for participation online/in social media (or other self-selection routes) rather than personal invitations<br>• “snowball” sampling (participants are asked to recruit further participants themselves)<br>(*Exception:* A special type of snowball sampling - called RDS or *respondent-driven sampling* - is appropriate when the targeted population is small or hard to identify and invite via traditional methods, like transgender people, people living |

| | |
|---|---|
| | with a rare disease, drug users etc.) <u>This needs to be specifically stated as such in the methods section.</u><br>● sampling from a specific subset of the population, which might differ from the target population, sampling from a particular occupational group/patient group/hobby group/living situation group not representative of the target population<br>***Example***: *target population being children in Italy, but the sample comes from two private schools, target population is elderly people and only retired teachers have been invited to the survey*<br>● quota sampling (i.e. sampling via a polling company/website involving individuals that are representative of a population, based on specific characteristics such as age and sex)<br>*(<u>Exception</u>: The use of a random sampling procedure is clearly described)*<br><br>✓ **Low risk** if at least one of the followings holds:<br>● the whole target population was invited to participate<br>● a randomly selected sample from the target population was used<br><br>? **Unclear risk**:<br>● when the method to invite participants and the specific environment and context where the sample was chosen from have not been specified *and when the target population is not defined* |
| **RoB Question 2:** How would you rate the risk of non-response bias? | This question is about *people who were invited and responded* to the study invitation by providing some data (not necessarily data for all study outcomes)<br><br>✗ **high risk** when at least one of the followings holds:<br>● There was a high percentage of non-responders (more than 30%)<br>● The methods of the survey suggest that non-response is likely to be the result of (typically negative) mental health status; for instance, people with very serious depression would not answer. As this is very difficult to judge, you can instead evaluate if non-response is likely to be linked with known predictors of mental health like sex, age, comorbidities or socioeconomical status.<br>***Example***: *If a questionnaire is administered to elderly via email rather than per post, it is less likely that participants of lower educational and professional status will answer the questionnaire, in this way possibly over- or underestimating the prevalence of depression.*<br><br>✓ **Low risk** when both of the following hold<br>● There was a low percentage of non-responders (equal or less than 30%)<br>● The methods of the survey suggests that non-responders probably have similar mental health status (or similar characteristics in sex, age, comorbidities and socioeconomical status) as responders |

| | |
|---|---|
| | ? **Unclear risk** when based on the information available a clear judgement is not possible. |
| **RoB Question 3:** How do you judge the risk of information bias? | This bias can be introduced when the instrument or method used<br><br>a) was not validated or when the outcomes were not assessed based on existing diagnostic criteria or generally the method to define the measured condition has a high risk to misclassify patients<br><br>b) is different between participants depending on their mental health status (or some probable mediator of mental health). If the instrument/method differs randomly between participants, this should not be considered a source of bias.<br><br>✘ **high risk** when any of the followings apply<br><br>● the instrument/method to record the condition is not validated (for the purpose of this review, we consider an instrument validated if clearly mentioned as such by the authors and references are provided).<br>● a trained investigator is required by the instrument but not used in the study<br>● the cut-off thresholds used in the study are not the ones identified by the instruments used<br>● the instrument/method to record the condition is different across groups of participants and these groups are not formed randomly.<br>***Example**: Self-answered BDI-II for people that felt well enough to have their appointment via video chat vs BDI-II filled together with the doctor for people that felt the need for an in-person visit at the clinic.*<br><br>✓ **Low risk** when all the following hold:<br><br>● the instruments used were validated (as mentioned by authors and documented by references)<br>● trained outcome assessors were used, if required by the instrument<br>● if thresholds are used for diagnosis, they are the ones commonly accepted for each instrument, as described in the study<br>● the instrument and method of assessing the condition is the same for all participants, or differences between participants are random.<br>**Example:** *All the participants were invited by e-mail; half were randomly selected to complete the survey themselves and half via video chat*<br><br>? **Unclear risk** when, based on the information available, a clear judgement is not possible. |
